# Serological Assays Estimate Highly Variable SARS-CoV-2 Neutralizing Antibody Activity in Recovered COVID19 Patients

**DOI:** 10.1101/2020.06.08.20124792

**Authors:** Larry L. Luchsinger, Brett Ransegnola, Daniel Jin, Frauke Muecksch, Yiska Weisblum, Weili Bao, Parakkal Jovvian George, Marilis Rodriguez, Nancy Tricoche, Fabian Schmidt, Chengjie Gao, Shabnam Jawahar, Mouli Pal, Emily Schnall, Huan Zhang, Donna Strauss, Karina Yazdanbakhsh, Christopher D. Hillyer, Paul D. Bieniasz, Theodora Hatziioannou

**Author notes:** Equal authorship. Co-corresponding authors # Address correspondence to Larry Luchsinger (Lead).

## Abstract

The development of neutralizing antibodies (nAb) against SARS-CoV-2, following infection or vaccination, is likely to be critical for the development of sufficient population immunity to drive cessation of the COVID19 pandemic. A large number of serologic tests, platforms and methodologies are being employed to determine seroprevalence in populations to select convalescent plasmas for therapeutic trials, and to guide policies about reopening. However, tests have substantial variability in sensitivity and specificity, and their ability to quantitatively predict levels of nAb is unknown. We collected 370 unique donors enrolled in the New York Blood Center Convalescent Plasma Program between April and May of 2020. We measured levels of antibodies in convalescent plasma using commercially available SARS-CoV-2 detection tests and in-house ELISA assays and correlated serological measurements with nAb activity measured using pseudotyped virus particles, which offer the most informative assessment of antiviral activity of patient sera against viral infection. Our data show that a large proportion of convalescent plasma samples have modest antibody levels and that commercially available tests have varying degrees of accuracy in predicting nAb activity. We found the Ortho Anti-SARS-CoV-2 Total Ig and IgG high throughput serological assays (HTSAs), as well as the Abbott SARS-CoV-2 IgG assay, quantify levels of antibodies that strongly correlate with nAb assays and are consistent with gold-standard ELISA assay results. These findings provide immediate clinical relevance to serology results that can be equated to nAb activity and could serve as a valuable ‘roadmap’ to guide the choice and interpretation of serological tests for SARS-CoV-2.

## Introduction

In late 2019, a cluster of patients in Wuhan, the capital city of China’s Hubei providence, were reported to be afflicted with a severe respiratory illness of unknown origin.(1, 2) Patients presented with symptoms that included high fever, pneumonia, dyspnea, and respiratory failure. The causative agent was identified to be severe acute respiratory syndrome coronavirus variant 2 (SARS-CoV-2), the 7^th^ coronavirus strain to infect humans to date,(3) and the clinical syndrome was designated coronavirus disease of 2019 (COVID19). The pathogenesis of COVID19 is similar to previously documented respiratory distress syndromes caused by related coronaviruses, including the 2005 SARS coronavirus (SARS-CoV) and the middle east respiratory syndrome coronavirus (MERS).(4) However, the greater transmissibility of SARS-CoV-2 has enabled a swift global spread that has resulted in substantial mortality. Detection and tracking SARS-CoV-2 spread has been difficult. Moreover, the spectrum of symptomatology observed in SARS-CoV-2 infection is wide, ranging from asymptomatic and mild, reminiscent of numerous seasonal infections, including influenza and common cold viruses, all the way to life-threatening respiratory failure that requires intensive care and invasive ventilation. Currently, increased age and comorbidities are the factors most highly predictive of severe of COVID19 disease.(5)

The utility of serological tests to identify individuals who have acquired antibodies against SARS-CoV-2 is thus recognized as both an indication of the seroprevalence of SARS-CoV-2 infection and, potentially, of immunity afforded to the seropositive individual.(3, 6-8) Seroconversion is determined by detection of antibodies that recognize SARS-CoV-2 antigens. Coronaviruses have 4 major structural proteins: spike (S) protein (including the S1 protein and receptor binding domain (RBD)), nucleocapsid (N) protein, membrane (M) protein and envelope (E) protein.(9) Previous studies of SARS-CoV and MERS found the most immunogenic antigens are the S- and N-proteins,(10) and development of serological tests for SARS-CoV-2 antibodies has focused heavily on these viral proteins.

Three major platforms of serological testing have been adopted; 1) enzyme linked immunosorbent assays (ELISA), 2) high-throughput serological assays (HTSA), and 3) lateral flow assays (LFA). ELISAs offer wide flexibility for research laboratories to select virtually any antigen of interest and provide highly sensitive, quantitative results. HTSAs are more suitable for clinical laboratories and offer limited antigen diversity but allow high-throughput and sensitive, semi-quantitative results. LFAs also offer limited antigen diversity, but function with small volumes (~20μL) of whole blood, plasma or sera and allow rapid (≤15 minutes) results at the point of care. The clinical community will undoubtedly employ multiple SARS-CoV-2 serology platforms but a comparative analysis across platforms has not been undertaken. Further, it is currently unknown whether the detection of antibodies that bind these proteins predicts neutralizing activity or protection against infection.(11)

Convalescent plasma (CP) transfusion has been recognized as a potential treatment for critically ill COVID19 patients and the New York Blood Center (NYBC) has led the first COVID19 CP donation program in the United States. Using 370 unique CP donor samples deposited in our COVID19 Research Repository (https://nybc.org/covid19repository), we conducted ELISA, HSTA and LFA assays as well as SARS-CoV-2 pseudovirus neutralization assays. We find that CP donors have a wide range of antibody titers measured across multiple COVID19 serological and neutralization assays. Notably, we show that some HTSA and ELISA assays predict neutralizing activity *in vitro* and may thus serve to predict antiviral activity against SARS-CoV-2 *in vivo*.

## Results

### Characteristics of the NYC CP Donor Population

Serological analysis of the CP donors was performed using 370 unique samples collected between April and May of 2020 from the NYC area. CP donors enrolled in the program were required to have tested positive for SARS-CoV-2 by PCR diagnostic tests and be symptom free for at least 2 weeks. To profile CP donors, we cross-referenced donor demographic data to the 2010 U.S. Census database.(12) CP donors had a median age of 41 years (95% CI: 39-44, range 17-75 years,) and showed a gaussian distribution (n=183, r^2^=0.89) compared to the national median age of 38.2 years in 2018 (**Figure 1A**). The frequency of male and female CP donors was 45.2% and 54.8%, respectively, and was not statistically different from the national average of 49.2% and 50.8% (**Figure 1B**). The frequency of ABORh blood group antigens was also largely consistent with the national frequency, with a slightly higher number of A^-^ and O^-^ donors and slightly lower number of AB+ and B+ donors than expected (**Figure 1C**). Finally, CP donor ethnicity was largely consistent with the national ethnic composition, with a slightly higher number of multiracial/other donors and lower number of Black/African American donors than expected (**Figure 1D**). Overall, the composition of NYC CP donors analyzed was reflective of the United States population demographic.

**Figure 1:**
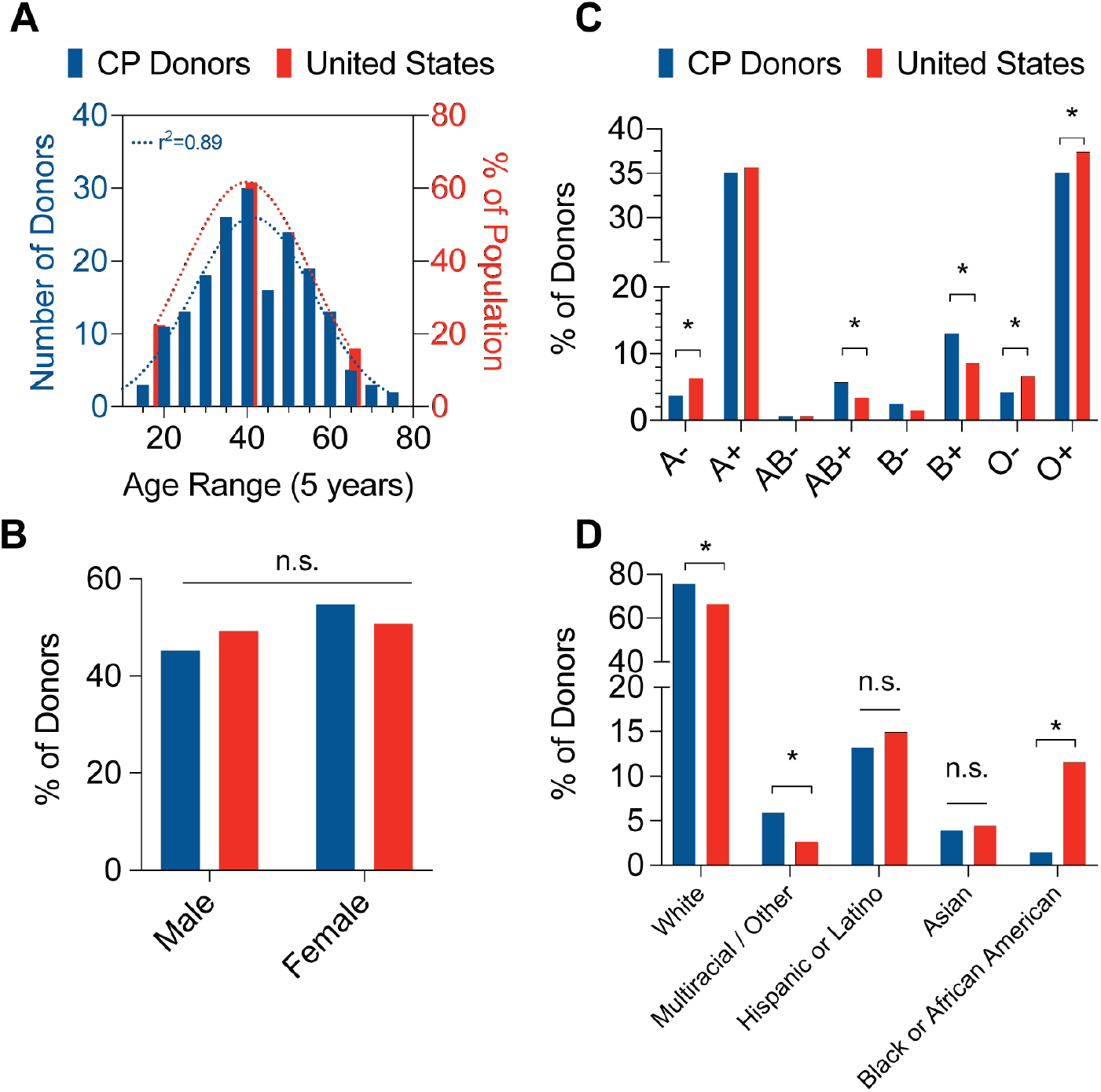
Demographics of convalescent plasma donors. **A;** Distribution of convalescent plasma donor age (left, blue bars) compared to U.S. population (right, red bars). Dotted line represents Gaussian distribution curve fit. N=263; Pearson’s correlation coefficient. **B;** Distribution of convalescent plasma donor blood group antigen (left, blue bars) compared to U.S. population (right, red bars). N=370, binomial test for discrepancy versus U.S. population; * p < 0.05. **C;** Distribution of convalescent plasma donor sex (blue bars) compared to U.S. population (red bars). N=354,binomial test for discrepancy versus U.S. population. **D;** Distribution of convalescent plasma donor ethnicity (blue bars) compared to U.S. population (right, red bars). N=204, binomial test for discrepancy versus U.S. population; * p<0.05.

### Neutralizing Activity of the CP Donor Population

Neutralization assays measure how effectively donor plasma or serum can inhibit virus infection of target cells and are the gold standard for measuring the antiviral activity of antibodies. In the case of SARS-CoV-2, such assays require in biosafety level 3 (BSL-3) facilities and highly trained personnel. To overcome this limitation and expedite testing, we employed pseudotyped virus assays based on either HIV-1 (human immunodeficiency virus type 1) or VSV (vesicular stomatitis virus). Both viruses were engineered to lack their own envelope glycoproteins and to express a luciferase reporter gene. Complementation *in trans* with the SARS-CoV-2 Spike (S) protein results in the generation of pseudotyped virus particles that are dependent on the interaction between the S protein and its receptor ACE2 (angiotensin-converting enzyme 2) for entry into cells.(13) These reporter viruses were used to measure infection of human cells engineered to express ACE2 (HIV-S assay) or expressed endogenous ACE2 (VSV-S assay) and to determine the ability of plasma dilutions to inhibit S-dependent virus entry. The NT_50_ values, reflecting the plasma dilution at which virus infection is reduced by 50%, were calculated for each sample (**Supplementary Figure 1A**).

The neutralizing activity of CP donor samples was extremely variable and NT_50_ values obtained ranged from <50 to over 20,000. The median NT_50_ values were 390.1 (95% CI: 278.3-499.7) and 450.6 (95% CI: 367.7-538.4) for the HIV-S or VSV-S assays, respectively (**Figure 2A**) and the two assays showed a high degree of correlation (**Supplementary Figure 1B-C**). Fresh frozen plasma (FFP) samples donated in 2019, before the SARS-CoV-2 outbreak, were used as negative controls (n=10). Importantly, the NT_50_ values of all FFP samples were ≤50, which is the highest concentration of plasma used in the neutralization assays and is hence designated as the signal cutoff (S/co) value. Overall, 83.1% and 92.7% of the CP donor samples had detectable neutralization activity using HIV-S and VSV-S assays, respectively (**Figure 2B**). Notably, 11.2% and 8.7% of CP donors had NT_50_ values at or greater than 2000 (40-fold over S/co) using HIV-S and VSV-S, assays respectively while 55.8% and 52% of CP donors had NT_50_ values at or less than 500 (10-fold over S/co) (**Figure 2B**). Thus, the majority of CP donors may have relatively modest neutralizing activity and a small proportion of donors have high neutralization activity.

**Figure 2:**
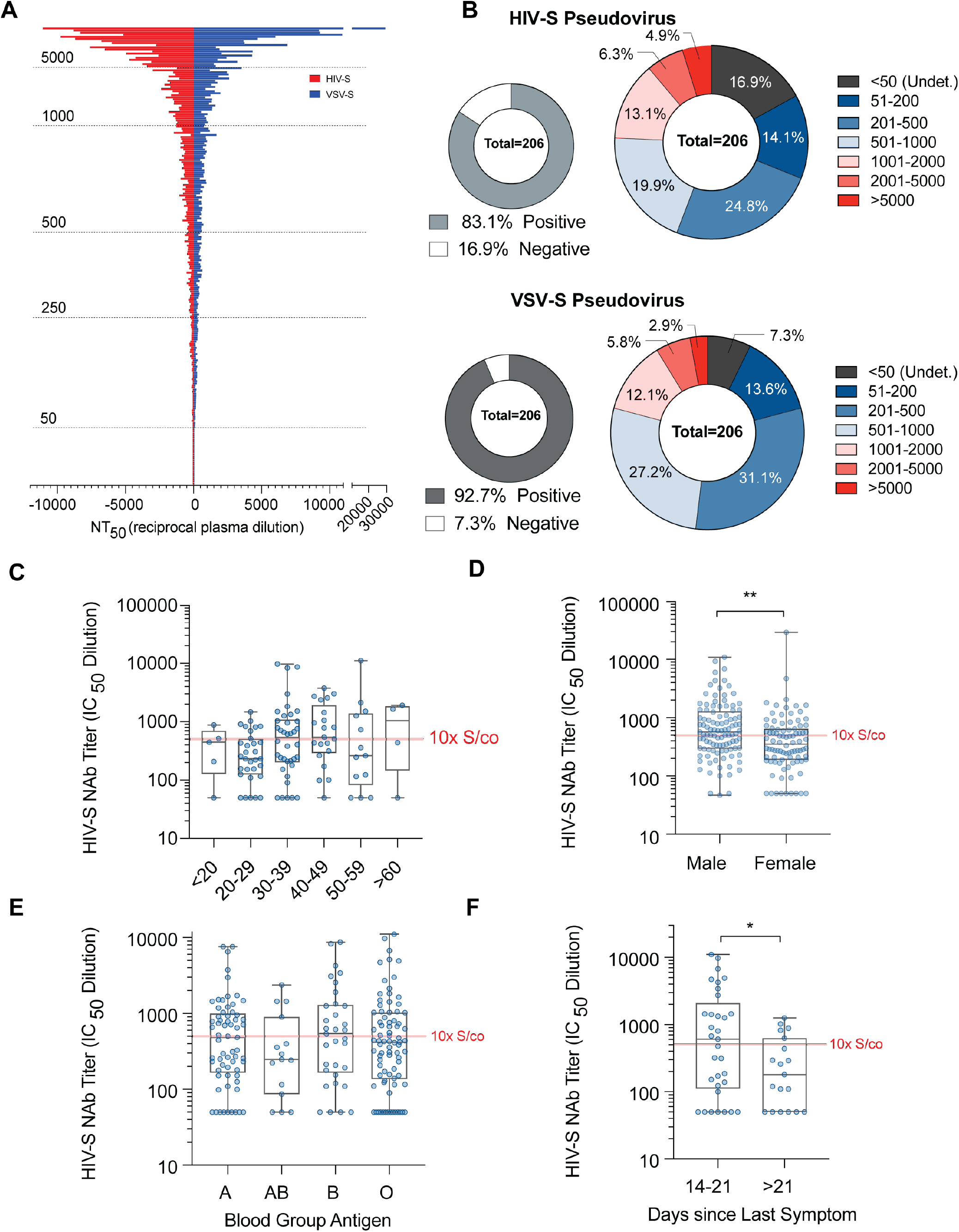
Neutralizing activity analysis of convalescent plasma donors. **A;** Distribution of neutralization IC_50_ values (NT_50_, reciprocal plasma dilution) of convalescent donor plasma using HIV (red) or VSV pseudovirus (blue) overexpressing the SARS-CoV-2 spike protein (S). **B;** Frequency of convalescent plasma donor NT_50_ values within indicated groups using HIV-S (top) or VSVS pseudovirus constructs. **C;** Frequency distribution of convalescent plasma HIV-S NT_50_ values versus age groups. Signal to cutoff (S/co, dotted grey line) and 10x S/co (solid grey line) thresholds are indicated. n=5-38, Kruskal-Wallis test; * p < 0.05. **D;** Frequency of convalescent plasma donor NT_50_ values versus sex. Signal to cutoff (S/co, dotted grey line) and 10x S/co (solid grey line) thresholds are indicated. n=190, Mann-Whitney test, ** p < 0.01. **E;** Frequency of convalescent plasma donor NT_50_ values versus blood group antigen. Signal to cutoff (S/co,dotted grey line) and 10x S/co (solid grey line) thresholds are indicated. n=15-82, Kruskal-Wallis test, * p < 0.05. **F;** Frequency of convalescent plasma donor NT_50_ values versus time (days) since last reported symptom. Signal to cutoff (S/co, dotted grey line) and 10x S/co (solid grey line) thresholds are indicated. n=19-33, Mann-Whitney t-test, *p < 0.05.

NT_50_ values were not statistically different between blood groups (**Figure 2E, Supplementary Figure 1G**) or age groups (**Figure 2C, Supplementary Figure 1E**) and there was no linear correlation of NT_50_ values with age (**Supplementary Figure 1D**) in contrast to previous reports.(14) However, in agreement with recent studies,(15) NT_50_ values of male CP donor samples were ~1.7-fold higher than those from female CP donors using HIV-S and VSV-S assays (**Figure 2D and Supplementary Figure 1F**, n = 195, p = 0.009 and <0.001, median difference 217 and 197, respectively). For CP donors where symptom dates were reported, the time between last symptom and the date of donation was calculated. Interestingly, CP donors 2-3 weeks post symptoms had a statistically significant increase in NT_50_ values compared to CP donors >3 weeks post-symptom (**Figure 2F and Supplementary Figure 1H**, n=52, p = 0.03 and 0.04, median difference 426 and 226, respectively). Overall, these data suggest CP donors possess a wide range of neutralizing antibody levels that are proportionately distributed across demographic categories with the exception of a small sex-dependent effect.

### Serological Test Results of the CP Donor Population

Multiple platforms have been deployed to detect seroconversion against SARS-CoV-2. The simplest tests are LFAs, which solubilize antibodies from whole blood, plasma or sera in an aqueous mobile phase which moves across a nitrocellulose membrane coated with anti-human IgG and/or IgM to distinguish between specific classes of immunoglobins while a control band ensures test function. Binding of antibodies to antigen-conjugated enzyme, such as horseradish peroxidase, generates a colored band at the test lines. Analysis of 144 CP donor samples showed that only 79.4% of CP donors tested positive for SARS-CoV-2-specfic IgG antibodies and 24.8% for IgM antibodies (**Figure 3A, top**). While LFAs are not designed to perform quantitatively, large discrepancies in band intensity between donors (**Supplementary Figure 2A**) is often presumed to indicate semi-quantitative results. We performed densitometric analysis of the test bands from LFA cassettes (**Supplementary Figure 2B, 2C**) and normalized each test to control band intensity. LFAs showed an intensity range of 0% – 99.2% for IgG bands and 0% – 18.5% for IgM bands, with a median intensity of 20% for IgG and <1% for IgM (**Figure 3A, bottom**). Thus, LFAs have a high degree of variation in band intensity within the CP donor population.

**Figure 3:**
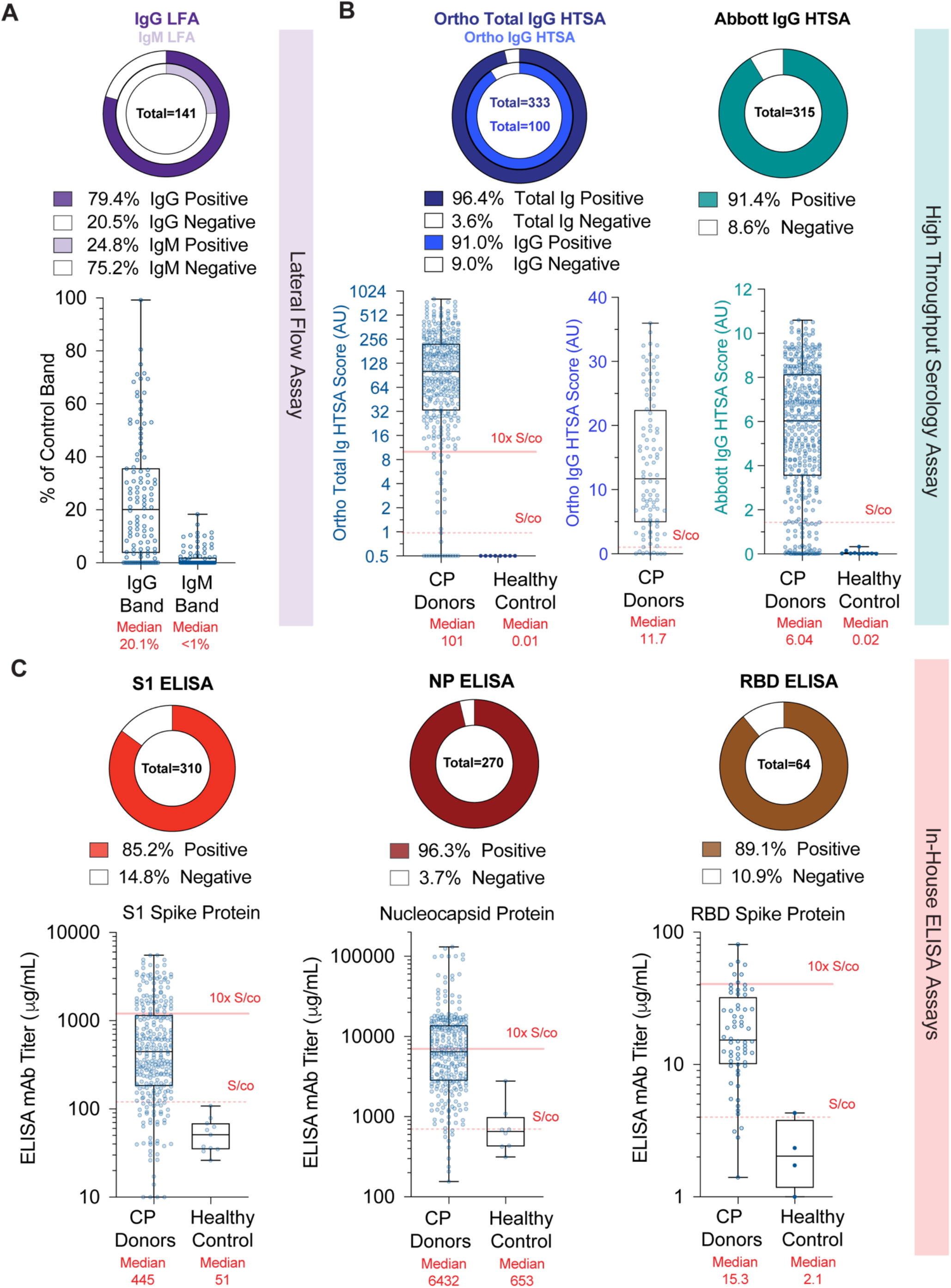
Serological analysis 592 of convalescent plasma donors. **A;** Frequency of densitometric IgG (left) or IgM (right) results from LFA bands relative to control band. Median values (red band) with 1^st^ and 3^rd^ quartiles (thin red lines) are shown. **B;** Frequency of HTSA results using the total Ig or IgG assays derived from the Ortho Diagnostics platform (left) or Abbott IgG assay platform (right). Results from fresh frozen plasma (FFP) units collected before COVID19 are shown as healthy controls. **C;** Frequency of S1 spike protein (left), Nucleocapsid (NP) protein (center) and RBD spike protein (right) ELISA titer results. Titers reflect concentrations calculated using a mAb standard curve and not absolute plasma concentrations. Median values (red band) with 1^st^ and 3^rd^ quartiles (thin red lines) are shown.

HTSA systems offer the advantage of performing semi-quantitative seroconversion assays using clinical laboratory testing infrastructure at large scale. We performed the Ortho-Clinical Diagnostics VITROS SARS-CoV-2 total Ig assay, the VITROS SARS-CoV-2 IgG assay and the Abbott Labs Architect SARS-CoV-2 IgG assay using between 100 and 330 CP donor plasma samples. We found 96.4% and 91.0% of CP donor samples were positive using the Ortho total Ig and IgG assays, respectively, and 91.4% were positive using the Abbott IgG assay (**Figure 3B**). The median value of CP samples using the Ortho total Ig assay was 101 arbitrary units (AU) (n=333, 95% CI: 78.5 – 123, S/co = 1, range 0 to 1000 AU) while that of FFP healthy controls was 0.01 AU (n=8, 95% CI: 0.01 – 0.02). Similarly, the median value of CP samples using the Ortho IgG assay was 11.7 AU (n=100, 95% CI: 8.3 – 16.07, S/co = 1, range 0 to ~30 AU). For the Abbott assay, the median value of CP samples was 6.04 AU (n=315, 95% CI: 5.48 – 6.44, S/co = 1.4, range 0 to ~10 AU) while that of FFP healthy controls was 0.02 AU (95% CI: 0.01 – 0.15). These results clearly show HTSA platforms detect a wide variation in antibody levels in the CP donor population and offer greater dynamic range than LFA assays.

The gold standard for quantification of antigen-specific antibodies is ELISA assays. Studies of antibody responses during SARS-CoV and MERS outbreaks identified the S- and N-proteins as the dominant antigens. Therefore, we designed three indirect ELISA assays using SARS-CoV-2 recombinant, His-tagged, spike protein S1 domain (S1), spike protein RBD domain (RBD) and nucleocapsid protein (N). We utilized monoclonal antibodies demonstrated to bind antigen in a dose-dependent manner to generate standard curves from which antibody concentrations were calculated and FFP from healthy controls to determine signal cutoffs. Thus, we report our ELISA results as monoclonal antibody (mAb) titers. These ELISA assays showed that 85.2%, 89.1%, and 96.3% of CP donor samples were positive for antibodies against S1, RBD and N antigens, respectively (**Figure 3C**). Using the S1 ELISA, the median value for CP donor samples was 445μg/mL (n=285, 95% CI: 342 – 536μ,g/mL, S/co = 120μg/mL) and for FFP controls 100.9μ,g/mL (n=10, 95% CI: 78 – 120μ,g/mL). In the NP ELISA the median value for CP donor samples was 6432μg/mL (n=271, 95% CI: 2811 – 13792μg/mL, S/co = 700μg/mL) while in the RBD ELISA the median value of CP donor samples was 15.6μg/mL (n=43, 95% CI: 12.55 – 25.6μ,g/mL, S/co = 4μ,g/mL). Notably, the range of S1 and NP-binding antibody concentrations observed in the ELISAs was extreme, constituting a 1,000-fold difference in titers within the CP donor population. Taken together, these data demonstrate that CP donors have a wide range of concentrations of antibodies specific to immunogenic SARS-CoV-2 antigens, as measured across multiple serological platforms.

### Correlation of Serology Tests with Neutralizing Activity

It is not logistically feasible to implement neutralization assays as a measurement of antiviral antibodies at a scale of the general population. While quantification of seroconversion is practiced, controlled studies that determine the relationship between quantitative SARS-CoV-2 serology test results and neutralizing activity is sparse. We examined the correlation between serology and neutralization assays in the CP donor samples (**Figure 4A, Supplementary Figure 3A, Supplementary Figure 4C**). As expected, S1 ELISA titers showed a positive linear regression with NT_50_ values (r^2^ = 0.35) while the RBD ELISA titers showed slightly higher linearity (r^2^ = 0.38), commensurate with the fact that the RBD is a key target for neutralizing antibodies. Conversely, NP ELISA titers showed a comparatively lower degree of linear regression with neutralization activity (r^2^ = 0.09). By comparison, both the Ortho HTSA total Ig assay and the IgG assay showed higher (r^2^ = 0.45 for both) while the Abbott HTSA IgG assay showed lower linear regression with neutralization activity (r^2^ = 0.24). Although Ortho HTSAs and the Abbott HTSA IgG platforms quantify antibodies against S1 and NP antigens, respectively, a linear regression of r^2^=0.33 was calculated between these two HTSAs (**Supplementary Figure 3B**). As expected, linear regression between the Ortho total Ig and IgG assay was strong (r^2^ = 0.72) since the two assays measure the same epitope. LFA IgG densitometry measurements showed the poorest correlation with neutralization activity (r^2^ = 0.22).

**Figure 4:**
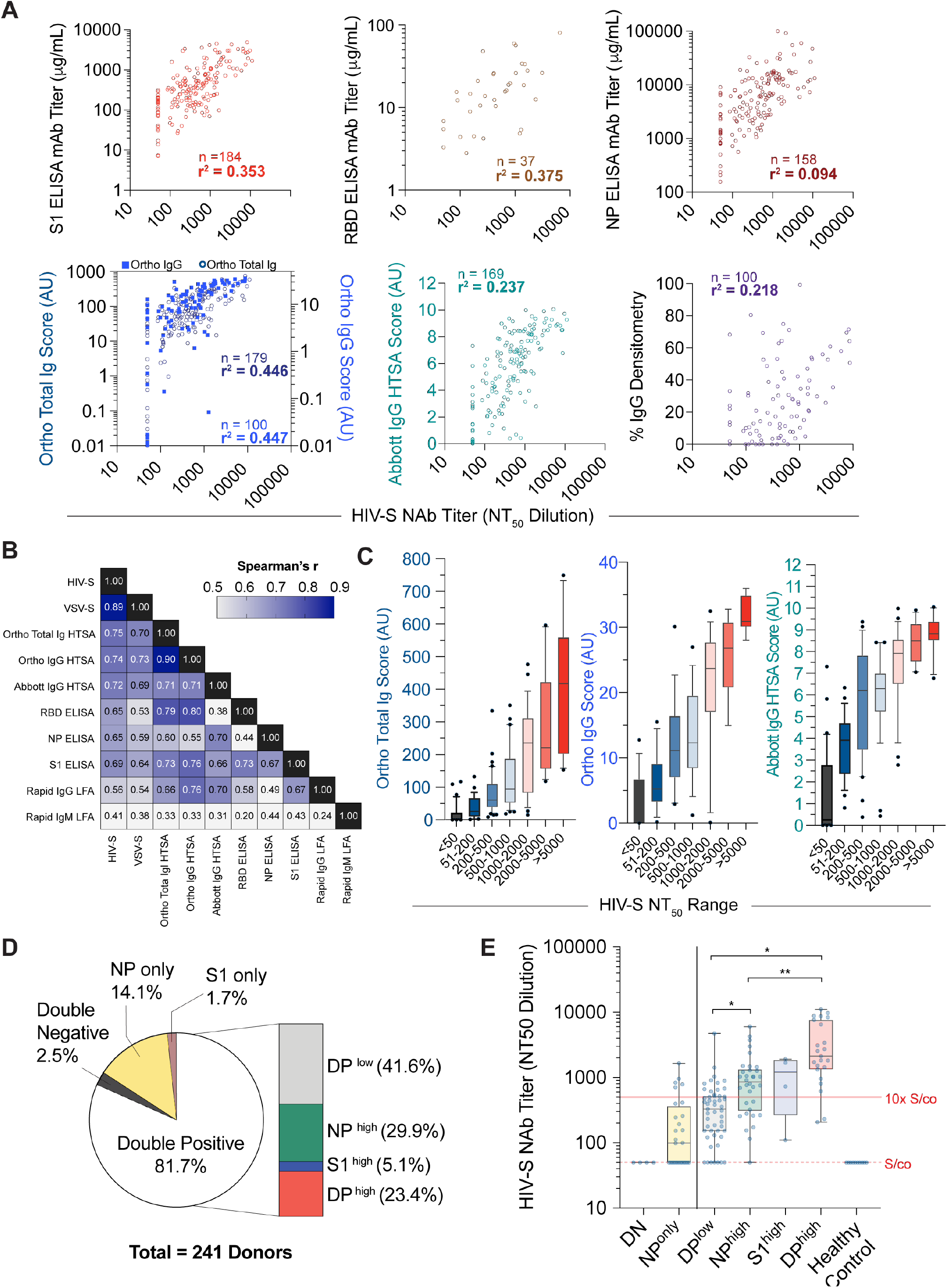
Correlation of serology assays versus neutralization activity of convalescent plasma donors. **A;** Linear regression of HIV-S NT_50_ values (abscissa) versus serological assay values (ordinate). N indicated in each graph, r_2_ = goodness of fit. **B;** Spearman correlation coefficients, *r*, of neutralization and serological assays. N=137 samples. **C;** Distribution of CP donor sample HTSA scores within indicated HIV-S NT_50_ groups using Ortho total Ig (left), Ortho IgG (center) or Abbott IgG (right) assays. **D;** Frequency of convalescent donor S1 and NP ELISA values defined in **C**. n=241 samples. **E;** Distribution of NT_50_ values corresponding to populations defined in **C**. n=4-51, Kruskall-Wallis test, * p < 0.05, ** p < 0.01.

Correlation between serological results and neutralization activity was also examined using the non-parametric Spearman test that does not assume linear dependence (**Figure 4B**). As expected, a high correlation between the HIV-S and VSV-S neutralization assays was obtained (r=0.89). The Ortho and Abbott HTSA platforms exhibited the highest degree of correlation with neutralization among the serology assays tested (r = 0.75 and 0.72, respectively for the HIV-S assay; 0.70 and 0.69 for the VSV-S assay). The S1, RBD, and NP ELISAs also showed a high degree of correlation, particularly with the HIV-S neutralization assay (r = 0.69, 0.65, and 0.65) while the LFA IgG and IgM assay showed the poorest correlation (r = 0.56, 0.41). Taken together, the data demonstrate that all quantitative serological assays correlate to some degree with neutralization activity. However, HTSA and S1 ELISA assays that measure anti-spike protein antigens have the highest predictive value as a surrogate for pseudovirus neutralization assays. Importantly, correlation between HTSA scores and NT_50_ values suggest presumptive ranges of neutralizing activity based on ranges of HTSA values (**Figure 4C, Supplementary Figure 4A**).

While ELISA assays revealed S1 and N antibody titers correlated with each other, these titers were not always proportional among CP donor samples. To examine the coincidence of S1 and NP antibody titers and using FFP plasma samples as negative controls, we categorized S1 and N antibody titers that fell below S/co values as ‘negative’ and titers greater 10-fold over S/co as ‘high’ (**Supplementary Figure 4B**). Using 241 CP donor samples that were assayed with both the S1 and N ELISA assays, we found that 81% of donors were double positive (DP), while 16% of samples were single positive (14% N and 2% S1, respectively) (**Figure 4D**). Only 2.5% of CP donors were double negative for S1 and NP antibodies. Within the double positive population, we found that 23% of samples were DP^high^ while 5% and 30% of samples were only S1^high^ or N^high^ and the remaining 42% were DP^low^. We then examined the distribution of NT_50_ values from the HIV-S neutralization assay within these populations (**Figure 4E**). Notably, DN samples showed NT_50_ values at the S/co observed for FFP healthy control samples while DP^low^ samples had relatively low NT_50_ values (median value 327, 95% CI: 186 – 444). Importantly, the DP^high^ donors had NT_50_ values that were 7-fold higher than DP^low^ donors (median value 2130). Additionally, NT_50_ values in the N^high^ and S1^high^ groups were 2.5- and 4-fold higher than those of the DP^low^ group.

Finally, we sought to determine if the frequency of peripheral blood immune cells varied as a function of antibody titer. We stained peripheral blood mononuclear cells (PBMCs) isolated from CP donor buffy coats for classical surface markers associated with B-cell or T-cell populations (**Supplementary Figure 5A, 5B**). We examined T cell subsets including T central memory (CD45RO+CD62L+) and T effector memory (TEM; CD45RO+CD62L^neg^) while B cell (CD20+) subsets analyzed included memory B cells (CD27+CD24+), plasmablasts (CD24^neg^CD38^hi^CD138^neg^) and the more mature plasma cells (CD24^neg^CD38^hi^CD138+) (**Supplementary Figure 5C**). We found statistically significant differences in naäve CD4 and CD8 T-cell populations in donors with high S1 ELISA titers compared to those with low titer. Decrease in CD24+CD27+ memory B cells was detected in individuals with higher anti-S titers. Although the cause of this lower frequency is not known, it could raise the possibility that individuals with reduced memory B cells may develop a less robust antibody response with future infections. Although our phenotypic analysis of B and T cell compartments was limited, these data suggest phenotypic differences in canonical B and T cell populations are insufficient to explain the large differences in antibody titers or neutralization activity observed in CP donors and warrants future studies designed to study B and T cell function from individual donors.

## Discussion

### Demographic limitations of the CP donor population

Recent studies have noted a disproportion in COVID19 morbidity and mortality among minority communities.(16) In this study, of the 370 CP samples analyzed, only 204 donors (55%) elected to identify ethnicity, representing the least reported demographic category we collected. Nevertheless, we did not observe a significant difference in nAb or serology results as a function of any demographic metric, including ethnicity. Although we showed that the CP donor samples analyzed in this study comprised a relatively normal distribution of demographic indicators, based on the U.S. census data, we acknowledge that some factors, including ethnicity, are underrepresented in this cohort and limit the interpretation of the study beyond the population aggregate. The potential explanations of this phenomenon are complex and extend beyond the scope of this study.(17) The blood banking community is continuously working to recruit minority donors, who are consistently underrepresented amongst regular blood donors.(18) Efforts to increase public participation in local blood and CP donor programs would both improve blood product diversity of transfusion products and strengthen the rigor of epidemiological disease. Thus, studies designed to characterize serological responses to COVID19 specifically in minority groups are warranted and necessary to augment our current understanding of the pandemic.

### Seroconversion assays of the population

Quantification of antiviral antibodies in recovered individuals is an important metric for determining population immunity conferred by exposure to SARS-CoV-2. Our study suggests that most New York City convalescent plasma donors have antibodies against SARS-CoV-2. Indeed, our data demonstrate that the HTSA, including Ortho and Abbot assays, which have received EUA from the FDA, are well suited to quantify a wide range of antibody titers and reported that 91 – 96.4% of the CP population possesses detectable SARS-CoV-2 antibodies. LFAs performed less well, and individuals with low antibody titers scored weakly positive or negative in LFAs. Such outcomes could be interpreted incorrectly, thus increasing the rate of false negative results. Ultimately, studies that accurately document SARS-CoV-2 seroprevalence in diverse populations will require highly sensitive, high quality assays such as HSTA or ELISA to be reliable.

### Correlation between serological assay measurements and neutralizing activity

Since patient recovery often precedes the development of efficacious and safe therapeutics, a longstanding treatment strategy for infectious diseases is passive antibody transfer. Therefore, refining strategies to improve CP infusion efficacy benefits both the current treatment options of COVID-19 and will inform the medical community for future pandemics. Our serological analyses are consistent with previous publications that show a considerable range in antibody titers in recovered COVID19 patients.(19) However, this study provides a comprehensive analysis of the correlation of quantitative serological test values with neutralization activity. Importantly, high dynamic range serological assays, such as the HTSA and S1 ELISA, had a significant linear correlation with neutralization activity. We show, for the first time, the extent to which three widely available SARS-CoV-2 HTSAs correlated to nAb activity as well as to each other, providing the clinical and scientific community with a comprehensive overview of clinical serology test performance. To this end, investigators from the Mayo Clinic’s COVID-19 Expanded Access Program (EAP) performed an exploratory analysis on the efficacy of CP as a therapeutic agent using data from over 35,000 transfusions.(20) Although the study showed uncertainty as to the statistical significance of effect, the authors noted that patients transfused with high antibody titer CP units, quantified by the Ortho IgG assay, showed a notable reduction in the odds ratio of mortality at both 7 and 30 days after transfusion. These data support the assertion that antibody quantification of CP units using high dynamic range HSTA assays may further improve therapeutic options for COVID-19 and, perhaps, future pandemic responses. This knowledge will also be necessary for deriving potential serologic correlates of protection,(21) and may aid in predicting immunity at the individual and population levels.(15)

Yet, the levels of plasma neutralizing activity required to prevent SARS-CoV-2 re-infection are currently unknown. Anecdotal results have been reported for seasonal coronavirus experimental infection studies. For example, one study of 229E HCoV found a positive correlation between pre-infection antibody titer and neutralization activity with symptom clinical severity.(22) In another study, 7 of 8 individuals with low neutralizing titers excreted virus upon re-exposure, compared to only 1 of 4 subjects with higher titers.(23) However, the conclusions of these studies are not directly comparable to the current SARS-CoV-2 pandemic. As such, the necessity of human epidemiological or vaccination studies are necessary to determine the minimum threshold of neutralizing activity necessary to prevent SARS-CoV-2 re-infection. Conversely, sub-neutralizing antibody levels have been reported to facilitate, rather than inhibit, viral entry of the some coronaviruses *in vitro*, through antibody dependent enhancement (ADE).(24-26) While ADE dependent replication has not been demonstrated to occur in SARS-CoV, viral uptake into macrophages via antibody association with Fc receptors does induce IL-6 and TNFα cytokines which may promote inflammation and tissue damage.(27) Insights gained from an accurate analysis of antibody levels and neutralization activity in SARS-CoV-2-infected individuals will help address these important questions and the corresponding health consequences.

A key biological question is: what underlies the large variation in antibody titers (neutralizing or otherwise) observed in CP donors? Numerous variables, including the effectiveness of innate immune responses, SARS-CoV-2 exposure dose, anatomical site of initial infection, and partial cross-reactive immunity conferred by prior seasonal coronavirus infection, could all impart variation on the amount and dissemination of SARS-CoV-2 antigen. Variation in the exposure of the adaptive immune system to SARS-CoV-2 antigen would, in turn, likely impact the magnitude of immune responses. Our observation that the levels of antibody to N, as well as S, correlates with S-specific neutralizing titer suggests that quantitative differences in the overall adaptive immune response to SARS-CoV-2, rather than intrinsic differences in the ability of individuals to mount neutralizing responses, at least partly explains the large variation in neutralizing capacity of CP. This notion is consistent with recent findings that all individuals examined, generated very similar, potent monoclonal SARS-CoV-2 neutralizing antibodies, but at very different levels.(15)

### Future utility for vaccine and CP donor strategies

The development of efficacious vaccines against SARS-CoV-2 may be necessary for ending the COVID19 pandemic. Clinical trials will undoubtedly include a battery of serological and neutralization assays in test subjects to assess candidate vaccine efficacy. Surrogate serology tests to neutralizing activity could help to rapidly inform as to the likely effectiveness, as well as immunogenicity, of vaccines against SARS-CoV-2. To this end, real-time analyses using scalable HTSA testing platforms is effectuate while future studies are conducted to more precisely measure *in vivo* neutralization activity.

Finally, the utility of convalescent plasma in the treatment of infection has been recognized since the turn of the 20^th^ century.(28) CP transfusion is thought to be effective through passive immunization, specifically the transfer of neutralizing antibodies from a recovered individual to another individual manifesting life-threatening symptoms.(29, 30) Previously CP therapy has been used to treat both SARS and MERS,(31) and currently can be rapidly deployed against SARS-CoV-2 while other therapies are under development.(32) Nevertheless, many questions remain regarding the optimal antibody levels necessary to treat patients at varying stages of COVID19 disease. Accurate quantification using serological assays that predict neutralization activity may improve clinical outcomes through refinement of CP unit selection for patients of varying symptomatology. In summary, we demonstrate that HTSA and S1 ELISA assays show the strongest correlation with neutralization activity and may serve to predict the degree of antiviral antibody activity present in recovered patients or vaccine recipients.

## Data Availability

Raw data will be made available upon request.

## Authors’ Contributions

LLL conceptualized the study, designed and performed serology experiments and managed the collection of data, figures and statistical analyses. LLL, PB, TH and CDH co-wrote the manuscript. TH, PB, FM, YW and FS designed and performed the neutralization assays. BR, DJ, WB, SJ, JP, MR and NT performed most of the serology experiments. CG, MP, ES and HZ processed and preserved donor plasma and PBMCs. DS coordinated donor demographic information. KY contributed to PBMC flow cytometry and interpretation. DJ and BR contributed equal authorship to the manuscript. LLL, PB and TH contributed equal corresponding authorship.

## Acknowledgements

We thank Jill Alberigo, Amanda Brites and Kelly Brightman from Rhode Island Blood Center for their help with performing the Ortho Anti-SARS-CoV-2 Total Test and the Abbott SARS-CoV-2 IgG test. We thank Chockalingam Palaniappan and Paul Contestable for their assistance with performing the Ortho Anti-SARS-CoV-2 IgG Test. We thank Haidee Chen for assistance with editing the manuscript.

## Methods

### Cell lines

Huh7.5 cells were a gift from Charles Rice(33). The 293T/ACE2cl.13 cell clone was generated by transducing 293T cells (ATCC® CRL-3216™) with a CSIB-based ACE2 lentivirus expression vector containing a cDNA encoding a catalytically inactive ACE2 mutant. Single cell clones were isolated by limiting dilution and one clone (293T/ACE2cl.13) was used in these studies.

### Collection of CP donor information, isolation of convalescent plasma and PBMCs

Disclosure of demographic information was elective at the time of donation and showed that of the 370 CP donors analyzed, 71.1% indicated age, 95.4% indicated blood type, 95.6% indicated sex and 55.1% indicated ethnicity. To examine the demographic characteristics within the convalescent plasma (CP) donor population, we used the 2010 U.S. Census demographic data as expected frequencies. Plasma was isolated from EDTA-anticoagulated human whole blood samples. Samples were shipped from the NYBC Sample Management Facility overnight at 4C and centrifuged for 5 min at 500 xg to facilitate plasma/cell phase separation. The resulting upper plasma layer was extracted, aliquoted to minimize future freeze-thaw cycles, and stored at −80 C. Samples were cryopreserved and stored in the NYBC COVID19 Research Repository (https://nybc.org/covid19repository).

### Plasmid constructs

The env-inactivated HIV-1 reporter construct (pHIV-1_NL4-3_ ΔEnv-NanoLuc) was generated from a pNL4-3 infectious molecular clone (obtained through NIH AIDS Reagent Program, Division of AIDS, NIAID, NIH from Dr Malcolm Martin). It contains a NanoLuc Luciferase reporter gene in place of nucleotides 1-100 of the nef-gene and a 940 bp deletion 3’ to the *vpu* stop-codon. The rVSVΔG/NG/NanoLuc plasmid was generated by insertion of a cassette containing an mNeonGreen/FMDV2A/NanoLuc luciferase cDNA into rVSVΔG (Kerafast) (PMID: 20709108) between the M and L genes. The pSARS-CoV-2 S-protein expression plasmid containing a C-terminally truncated SARS-CoV-2 S protein (pSARS-CoV2_Δ19_) was generated by insertion of a synthetic human-codon optimized cDNA encoding SARS-CoV-2 S1 spike protein lacking the C-terminal 19 codons into pCR3.1. An ACE2 lentiviral expression vector was constructed by inserting a cDNA encoding a catalytically inactive ACE2 mutant into the lentivirus expression vector CSIB (PMID: 30084827).

### SARS-CoV-2 pseudotype particles

To generate (HIV/NanoLuc)-SARS-CoV-2 pseudotype particles, 293T cells were transfected with pHIV-1_NL4-3_ ΔEnv-NanoLuc reporter virus plasmid and pSARS-CoV-2-S_Δ19_ at a molar plasmid ratio of 1:0.55. The transfected cells were washed twice with PBS the following day, and at 48h after transfection, supernatant was harvested, clarified by centrifugation, passed through a 0.22 μm filter, aliquoted and frozen at −80°C. To generate (VSV/NG/NanoLuc)-SARS-CoV-2 pseudotype particles, 293T cells were infected with recombinant T7-expressing vaccinia virus (vTF7-3) and transfected with rVSVΔG/NG/NanoLuc, pBS-N, pBS-P, pBS-L, and pBS-G (PMID: 20709108). At ~24h post transfection the supernatant was collected, filtered and used to infect 293T cells transfected with a VSV-G expression plasmid, for amplification. To prepare stocks of (VSV/NG/NanoLuc)-SARS-CoV-2 pseudotype particles, 293T cells were transfected with pSARS-CoV_Δ19_ and infected with the VSV-G complemented rVSVΔG/NG/NanoLuc virus. At 16h later the supernatant was collected, clarified by centrifugation, filtered, pelleted through a 20% sucrose cushion and stored at −80°C. The viral stock was incubated with 20% I1 hybridoma supernatant (ATCC CRL-2700) for 1h at 37°C before use.

### Neutralization assays

To measure neutralizing antibody activity in convalescent plasma, five-fold serial dilutions of plasma were incubated for 1 hour at 37°C in 96-well plates with an aliquot of HIV-1 or VSV-based SARS-CoV-2 pseudotyped virus containing approximately 1×10^3^ infectious units. Thereafter, 100 μl of the plasma/virus mixture was added to target cells (293T_Ace2_ cl.13, or Huh7.5) cells in 96-well plates. Cells were cultured for 48h (HIV-1 pseudotype viruses) or 16h (VSV pseudotype viruses). Then, cells were washed twice, lysed and NanoLuc Luciferase activity in lysates was measured using either the Nano-Glo Luciferase Assay System (Promega) and a Modulus II Microplate Multimode reader (Turner BioSystem) or a Glowmax Navigator luminometer (Promega). The half maximal neutralizing titer (NT_50_) for plasma, was determined using a 4-parameter nonlinear regression in Prism 8.4 (GraphPad).

### Lateral Flow ImmunoAssay (LFA)

Lateral flow immunoassays (LFAs) were provided by external companies. Assay cartridges contained detection bands for IgG and IgM against SARS-CoV2 specific epitopes as well as an internal positive control. For each assay, 20 μL convalescent plasma or serum was applied to the sample pad, followed by two drops of proprietary running buffer. After 30 minutes, high resolution pictures of the detection zone were taken and saved as.JPEG files. All tests were performed at room temperature.

### LFA Densitometry Analysis

Relative quantification of anti-SARS-CoV-2 IgG and IgM in convalescent plasma samples was performed using built-in gel analysis macros in FIJI (https://fiji.sc/). A rectangular selection covering the detection zone was analyzed using Analyze>Gels>Plot Lanes. Integrated density values were outlined manually and extracted from the resulting plot. Using MS Excel, IgG and IgM values were normalized against the density of the control band.

The remaining whole blood cellular phase was supplemented with 2 mL of 35 g/L HSA/DPBS and diluted 1:1 with DPBS. Diluted whole blood was layered over 7 mL Ficoll-Paque Premium 1.078 g/mL (GE Healthcare) and centrifuged for 20 minutes at 20C and 400xg without braking. Buffer coats were extracted, counted with AOPI viability stain using the Cellometer Auto2000 (Nexelom Bioscience LLC), and frozen in PBMC freezing media (10% DMSO in Knockout SR).

### SARS-CoV-2 Binding-Antibody ELISA

Flat-well, nickel-coated 96 well ELISA plates (Thermo Scientific) were coated with 2μg/mL of recombinant S1 spike protein, nucleocapsid protein, or Receptor Binding Domain (RBD) spike protein specific to SARS-CoV-2 in resuspension buffer (1% Human Serum Albumin in 0.01% PBST) and incubated in a stationary humidified chamber overnight at 4 C. On the day of the assay, plates were blocked for 30 min with ELISA blocking buffer (3% W/V non-fat milk in PBST). Standard curves for both S1 and RBD assays were generated by using mouse anti-SARS-CoV spike protein monoclonal antibody (clone [3A2], ABIN2452119, Antibodies-Online) as the standard. Anti-SARS-CoV-2 Nucleocapsid mouse monoclonal antibody (clone [7E1B], bsm-41414M, Bioss Antibodies) was used as a standard for nucleocapsid binding assays.

Monoclonal antibody standard curves and serial dilutions of convalescent donor plasma were prepared in assay buffer (1% non-fat milk in PBST) and added to blocked plates in technical duplicate for 1 hr with orbital shaking at room temperature. Plates were then washed three times with PBST and incubated for 1 hr with ELISA assay buffer containing Goat anti-Human IgA, IgG, IgM (Heavy & Light Chain) Antibody-HRP (Cat. No. ABIN100792, Antibodies-Online) and Goat anti-Mouse IgG2b (Heavy Chain) Antibody-HRP (Cat. No. ABIN376251, Antibodies-Online) at 1:30000 and 1:3000 dilutions, respectively. Plates were then washed three times, developed with Pierce TMB substrate for 5 min, and quenched with 3 M HCl. Absorbance readings were collected at 450 nm. Standard curves were constructed in Prism 8.4 (Graphpad Software Inc.) using a Sigmoidal 4PL Non-Linear Regression (curve fit) model.

### High-throughput Serology Assays

Convalescent donor plasma samples were barcoded and dispatched to Rhode Island Blood Center (RIBC). Samples were analyzed using the Abbott SARS-CoV-2 IgG chemiluminescent microparticle immunoassay with the Abbott Architect i2000SR (Abbott Core Laboratories), as well as the VITROS Immunodiagnostic Products Anti-SARS-CoV-2 Total Test and the Anti-SARS-CoV-2 IgG Test with the VITROS 5600 (Ortho Clinical Diagnostics). All assays were performed by trained RIBC employees according to the respective manufacturer standard procedures.

### Flow cytometric analysis of PBMCs

Cryopreserved PBMCs were thawed, filtered and stained with a B-cell or T-cell antibody cocktail for 30 minutes in PBS. Cells were washed with PBS and analyzed with a BD LSR Fortessa 4 laser cytometer. Cytometric analysis was performed using RUO FCS Express 7 (DeNovo Software).

